# Intragastric behavior of an experimental infant formula may better mimic intragastric behavior of human milk as compared to a control formula

**DOI:** 10.1101/2023.09.06.23295112

**Authors:** Julia J.M. Roelofs, Reina S. Tjoelker, Tim T. Lambers, Paul A.M. Smeets

## Abstract

**Background:** During breastfeeding the macronutrient composition of breastmilk changes gradually from relatively low-fat (foremilk) to relatively high-fat (hindmilk), initially exposing the gastrointestinal tract to a relatively low fat concentration. In contrast, infant formulae (IF) are homogenous. Mild processing and addition of milk fat globule membrane (MFGM) may impact gastric emulsion instability, potentially impacting the phased release of nutrients as observed during breastfeeding.

**Objective:** To assess gastric emulsion stability, gastric emptying, and the postprandial plasma metabolome of an experimental minimally processed IF (EF) with an altered fat-globule interface and a control IF (CF).

**Methods:** Twenty healthy males participated in this double-blind randomized crossover trial. Gastric MRI scans and blood samples were obtained before and after consumption of 600 ml CF or EF over a 2-h period. Outcomes included gastric top layer formation, total gastric volume, and blood parameters (FFA, insulin, glucose, and NMR-metabolomics).

**Results:** EF showed an earlier onset (13.4 min, p=0.017), smaller maximum volume (49.0 ml, p= 0.033), and a shorter time to maximum top layer volume (13.9 min, p=0.022), but similar AUC (p=0.915) compared to CF. Total gastric volume did not show a treatment*time effect. Insulin concentrations were lower for EF. FFA and glucose did not differ. EF yielded higher serum concentrations of phospholipid-and cholesterol-related metabolites.

**Conclusion:** An EF displayed faster gastric creaming than a CF, thereby potentially better mimicking the behavior of breastmilk which leads to phased release of nutrients into the intestine. Overall physiological benefits of this difference in gastric behavior remain to be studied further in infants.

## 1 Introduction

Breastmilk (BM) provides the best nutrition for the growth, development and health of infants (Victora et al., 2016; WHO, 2003). In case BM is not or not sufficiently available, or in case parents choose to feed their infant otherwise, infant formula (IF) is the only safe alternative to provide adequate nutrition (Dewey, 2003; WHO, 2003). Nevertheless, several reviews and meta-analyses report that differences in infant feeding may have different health effects. For instance, a longer duration of breastfeeding is associated with up to 26% reduced odds of overweight or obesity later in life (Harder et al., 2005; Victora et al., 2016; Weng et al., 2012).

One of the many differences between BM and IF is the change in fat content during a feed. During breastfeeding, the milk fat content and thus the caloric density of BM, slowly increases from low in foremilk, to relatively high in hindmilk (Daly et al., 1993; Forsum & Lonnerdal, 1979; Hytten, 1954; Italianer et al., 2020; Kent et al., 2006). With breastfeeding, the gastrointestinal tract will thus initially be exposed to a lower fat concentration. In addition, it is known that the fat content of BM varies greatly between individuals and shows diurnal variation (Khan et al., 2013). In contrast, the fat content of IF is stable during a feed and throughout the day. Physiologically, these differences may impact feed intake as e.g. the changes in fat content during breastfeeding are thought to influence satiation and overall feed intake (Perez-Escamilla et al., 1995).

Both BM and IF are essentially oil-in-water emulsions (Wang et al., 2023). The fat globules in BM are covered by a biological membrane called the milk fat globule membrane (MFGM). This membrane not only stabilizes the emulsion, but also facilitates digestion (Lopez, 2020). In contrast to BM, the fat globules in IF are primarily stabilized by proteins such as whey proteins and caseins (McCarthy et al., 2012). Under gastric conditions, as a result of pepsin-mediated protein digestion, these emulsions will destabilize, causing coalescence and creaming of the milk fat globules which results in a high-fat top layer (Kunz et al., 2005). Due to their different fat globule interface and coherent emulsion stabilities, BM and IF destabilize at different rates in the stomach (Bourlieu et al., 2015; Camps et al., 2021; Chai et al., 2022).

In addition to protein composition, formula processing also influences emulsion stability, mainly by heating-induced protein denaturation (Ye et al., 2020). Particularly whey proteins are sensitive to this and form complexes with caseins upon heating (Anema et al., 2004; Fox et al., 2015; Guyomarc’h et al., 2009; Mulet-Cabero et al., 2019). The formation of these complexes results in a higher emulsion stability because more whey proteins are absorbed at the fat-globule interface stabilized by caseins (Raikos, 2010). More severe heat treatment of IF may thus impact gastric emulsion stability resulting in slower coalescence and creaming in the stomach.

Overall, BM is known to have a faster gastric emptying rate compared to IF (Meyer et al., 2015; Van Den Driessche et al., 1999; Wang et al., 2023). This was also confirmed by Camps et al. (2021) with the use of Magnetic Resonance Imaging (MRI) in adults.

The advantage of using MRI in digestive research is that it allows for visualization and quantification of intragastric processes such as gastric emptying and emulsion stability (Smeets et al., 2021). However, due to ethical constraints, MRI cannot be used to assess gastric processes in healthy infants for research purposes.

Therefore, in this proof-of-concept study, gastric emulsion stability, gastric emptying, and post-prandial plasma metabolomics of an experimental IF, produced with a reduced heat-load and enriched in MFGM, was compared with a control IF in healthy adults. To complement this *in vivo* study, *in vitro* digestions were performed under both simulated infant and adult conditions to investigate emulsion stability of the formulae under gastric conditions. It was hypothesized that the different fat-globule interface resulting from differences in formula composition and processing would result in faster gastric emulsion instability thereby potentially better mimicking the phased-release of nutrients in the intestine as observed with breastfeeding.

## 2 Participants and methods

### 2.1 In vitro digestion

*In vitro* gastric digestion was performed using a semi-dynamic digestion model based on the international INFOGEST standard simulating infant and adult gastric conditions as described (Lambers et al., 2023). Digestion units contained 2.5 ml (infant), or 6 ml (adult) simulated gastric fluid (SGF), containing 30 mM HCl (Sigma-Aldrich) and 300 U/ml pepsin (Sigma-Aldrich, P6887) for infant or 100 mM HCl and 1000 U/ml pepsin for adult conditions, at the start of the experiment to simulate the fasting state. 60 ml formula was added immediately (adult) or with a feed flow of 3 ml/min (simulating a typical infant feeding time of 20 min) and SGF with a flow of 0.39 ml/min (infant) or 0.72 ml/min (adult) was added until sampling pH using preprogrammed DAS-box scripts. Subsequently, protease inhibitors (Pepstatin A, 5 µM, Sigma-Aldrich) were added to stop the enzymatic reactions and visually analyze layer formation over time. During the simulated digestion, samples were taken at pH values representing both the early and later phases of gastric digestion (Bourlieu et al., 2014) and layer formation was visually assessed with the use of photographs.

### 2.2 In vivo trial

#### 2.2.1 Design

This study was a double-blind randomized crossover trial in which healthy men underwent gastric MRI scans and blood sampling at baseline and after consumption of two formulae. The primary outcome was gastric top layer formation resulting from emulsion instability. Secondary outcomes were total gastric volume, and blood parameters related to metabolic responses, including free fatty acids (FFA), glucose, insulin, and a range of (Nuclear Magnetic Resonance (NMR)-based) metabolites. In addition, subjective ratings of appetite (hunger, fullness, thirst, desire to eat, and prospective consumption) and nausea were collected. The study procedures were approved by the Medical Ethical Committee of Wageningen University in accordance with the Helsinki Declaration of 1975 as revised in 2013. The study was registered with clinicaltrials.gov under number NCT05224947. All participants signed an informed consent.

#### 2.2.2 Participants

Twenty healthy (self-reported) males aged 18-45 y and with a BMI between 18.5 and 25 kg/m^2^ were included. Participants were excluded if they reported an allergy or intolerance for cow’s milk, lactose, soy and/or fish, gastric disorders, or regular gastric complaints, used medication that affects gastric behavior, smoked more than 2 cigarettes per week, had an alcohol intake >14 glasses per week, or had a contra-indication to MRI scanning (including but not limited to pacemakers and defibrillators, ferromagnetic implants, and claustrophobia). Since female sex hormone levels are known to influence gastrointestinal function, only males were included in the study (Gonenne et al., 2006; Lajterer et al., 2022; Soldin & Mattison, 2009). Participants were recruited via digital advertisements (e-mail and social media). In total, 20 men participated in the study (age: 25.5 ± 5.8 y, BMI: 21.9 ± 1.5 kg/m^2^) (**Supplementary Figure 1**).

#### 2.2.3 Treatments

A routine cow’s milk-based formula (control IF, CF) and an experimental mildly processed formula (i.e. low-temperature pasteurization) comprised of skimmed milk, a native whey protein concentrate (Hiprotal® Milkserum 60 Liquid, FrieslandCampina), and a MFGM-enriched whey protein concentrate (Vivinal® MFGM, FrieslandCampina) (experimental IF, EF) were used. Both products met the nutritional requirements of infants (0 to 6 months) (**Table 1**), were produced specifically for this study, and were produced under food-grade conditions. Participants consumed 600 ml of the formulae at approximately 37 °C as prescribed in the preparation instruction of both formulae.

**Table 1.** Nutrient composition of the two formulae.

| Composition per 100 ml | Control | Experimental |
| --- | --- | --- |
| Energy (kcal (kJ)) | 66 (276) | 67 (280) |
| Protein<sup>[N*6.25]</sup> (g) | 1.4 | 1.3 |
| - Casein<sup>[N*6.25]</sup> (g) | 0.5 | 0.5 |
| - Whey protein<sup>[N*6.25]</sup> (g) | 0.9 | 0.8 |
| Fat (g) | 3.5 | 3.4 |
| Carbohydrate (g) | 7.0 | 7.3 |
| Galacto-oligosaccharides (g) | 0.4 | 0.4 |

#### 2.2.4 Study procedures

Participants were instructed to consume the same meal the evening prior to both test days. After this the overnight fasting period of minimally 12 hours started. Drinking water was allowed up to 2 hours prior to their visit. Upon arrival at Hospital Gelderse Vallei (Ede, The Netherlands), a cannula was placed in an antecubital vein, a blood sample was taken, a baseline MRI scan was performed, and subjective ratings on appetite and nausea were obtained. Subsequently, participants consumed one of the two formulae from a cup while in an upright position. Mean (± SD) ingestion time was 2.1 (± 0.9) min (CF: 2.3 ± 0.9 min, EF: 2.0 ± 0.9 min).Subsequently, gastric MRI scans were performed at 5-minute intervals during the first 30 minutes after the start of consumption. After that, scans were made every 10 minutes, up until 2 hours. Blood samples were taken at t = 15, 30, 45, 60, 75, 90 and 120 min. In addition, participants were asked to verbally rate their appetite and nausea on a scale from 0 (not at all) to 100 (very much) every 10 minutes, up to 90 minutes (Noble et al., 2005) (**Figure 1**).

**Figure 1.**
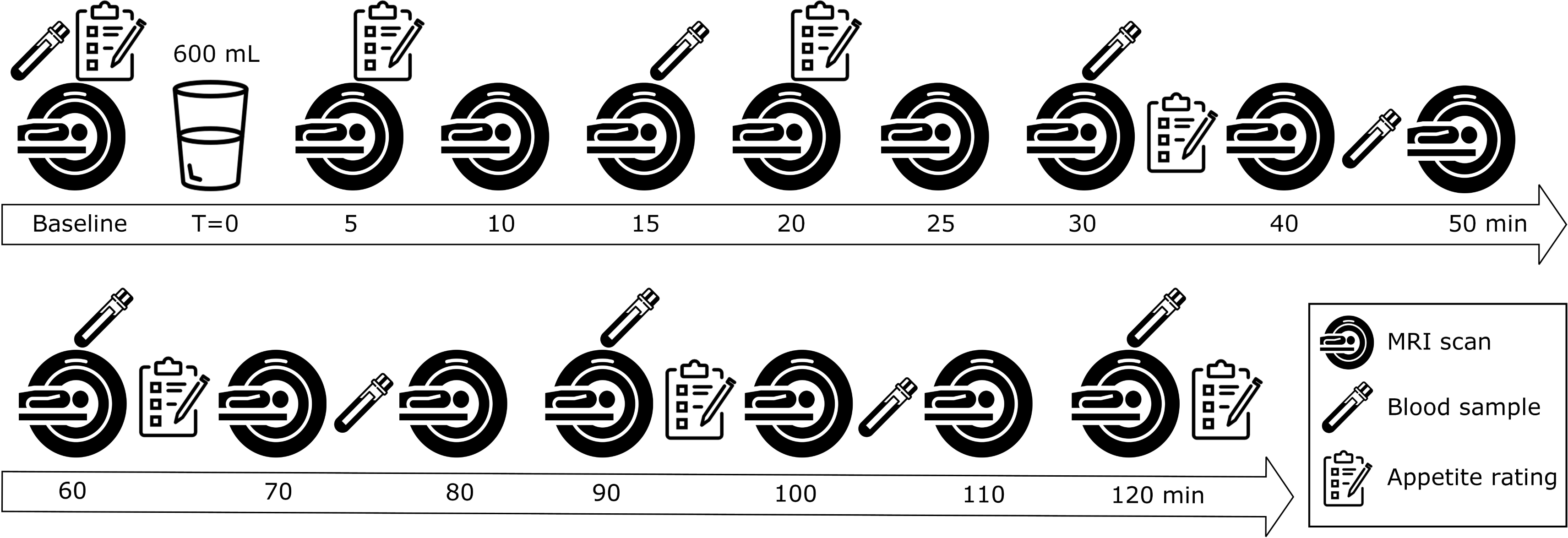
Overview of a test session.

#### 2.2.5 MRI scanning

Participants were scanned in a supine position with the use of a 3 Tesla Philips Ingenia Elition X MRI scanner (Philips, Eindhoven, The Netherlands). A 2-D Turbo Spin Echo sequence (37 4-mm slices, 1.4 mm gap, 1 x 1 mm in-plane resolution, TR: 550 ms, TE 80 ms, flip angle: 90 degrees) was used with breath hold command on expiration to fixate the position of the diaphragm and the stomach. The scan lasted approximately 20 seconds.

#### 2.2.6 MRI image analysis

Gastric top layer volume and total gastric content volume were manually delineated with the use of the program MIPAV (Medical Image Processing, Analysis and Visualization Version 7.4.0, 2016) (**Figure 2**). Top layer volume and total gastric volume were calculated for each time point with the use of MATLAB (2021b), taking into account slice thickness and gap distance. A representative example of a complete time series for both formulae is shown in the supplement (**Supplementary Figure 2**).

**Figure 2.**
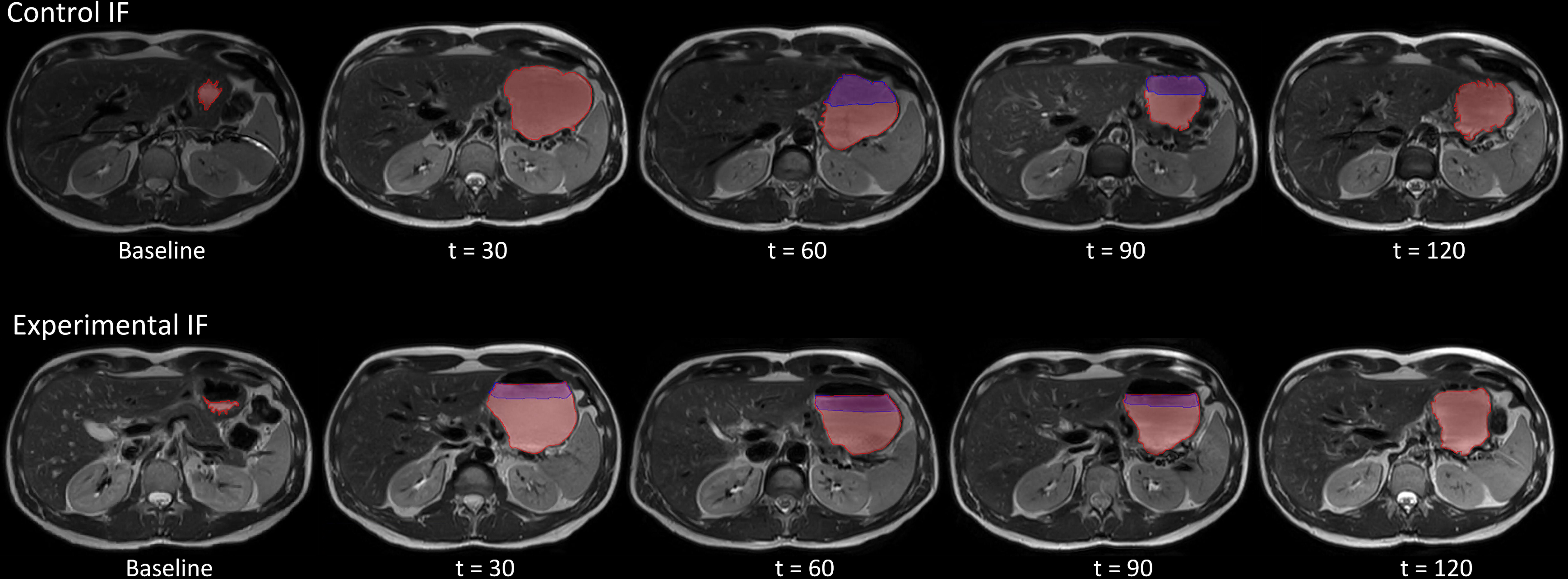
Illustration of a stomach MRI time series from one participant with top layer volume (blue) and total gastric volume (light red) delineated for the control IF and experimental IF.

The stomach contents were independently delineated by two researchers. To ensure that both researchers delineated total gastric volume and top layer in the same manner, the scans of both sessions of three participants were assessed by both researchers. The delineations were then discussed, and consensus agreements were made to ensure similar assessment by both observers. After this, scans were delineated by one of the two researchers. 15% of the sessions were analyzed in duplicate to check for potential differences. For top layer volume this resulted in an interclass correlation coefficient of 0.903 (95% CI: 0.849 – 0.938) which indicates good reliability. For total gastric content volume this was 0.997 (95% CI: 0.993-0.998), indicating excellent reliability (Koo & Li, 2016).

#### 2.2.7 Blood sample collection and analysis

Blood samples were drawn from the IV cannula into sodium-fluoride, serum-, and EDTA tubes. After collection, the serum tubes were allowed to clot for 60 min. All tubes were centrifuged at 1000 g for 10 min. The sodium-fluoride and serum tubes were centrifuged at 22°C and the EDTA tubes at 4°C. The aliquots were stored at −80°C until they were analyzed in bulk. To determine glucose concentrations, the sodium-fluoride plasma samples were processed using the Atellica CH Glucose Hexokinase_3 (GluH_3) assay kit and quantified using the Atellica CH analyzer (Siemens Healthineers, Netherlands) by a clinical chemistry laboratory (Ziekenhuis Gelderse Vallei, Ede, The Netherlands). The lower detection limit was 0.2 mmol/l and a maximum intra-assay CV of 4.5%. The EDTA plasma samples were processed and insulin was quantified using an enzymatic immunoassay kit (ELISA, Mercodia AB, Sweden). The lower detection limit was 1 mU/L and inter-assay CVs ranged from 3.3-19.1%. For the quantification of FFA, EDTA plasma samples were processed and quantified with an enzymatic kit (InstruChemie, Delfzijl, The Netherlands). The lower detection limit was 0.008 mmol/l and inter-assay CVs ranged from 0.2-20.9%. Quantification of 250 (NMR-based) metabolites was performed in the serum samples using ^1^H-NMR metabolomics (Nightingale Health Ltd, Helsinki, Finland, https://nightingalehealth.com/) (Würtz et al., 2017).

#### 2.2.8 Sample size

Sample size was calculated for the primary outcome top layer formation. The calculation was based on Camps et al. (2021) who found that an intake volume of 200 ml IF resulted in a top layer of 17 ± 2.3 ml in adults. Therefore, it was expected that intake of 600 ml IF would result in a top layer of approximately 50 ml. With a larger intake volume, the magnitude of individual differences was expected to increase. Thus, it was estimated that the deviation from control would increase to approximately 13 ml. The minimal MRI-detectable difference in top layer deemed clinically relevant was estimated at 10 ml. Based on a two-sided test, an α of 0.05 and a power of 0.9, it was estimated that 20 complete datasets were needed. The calculation was done using: http://hedwig.mgh.harvard.edu/sample_size/js/js_crossover_quant.html.

#### 2.2.9 Statistical analysis

For gastric top layer formation, the time at which the top layer appeared (onset) was compared between treatments. In addition, the maximum top layer volume and time to maximum top layer volume was identified for each individual. All three parameters were compared between treatments with a paired t-test and with a linear mixed model that included baseline gastric volume as covariate. AUC of the top layer and total gastric volume over time were also calculated for each individual using the trapezoidal rule and compared with paired t-tests. Pearson correlation coefficients were calculated for baseline gastric volume and top layer characteristics.

Differences in gastric top layer volume over time were tested with the use of a generalized linear mixed model using a zero-inflated Poisson distribution, testing for main effects of time, treatment, and treatment*time interactions. Baseline gastric volume was included as a covariate, due to its effect on digestion (Camps et al., 2021). Differences in total gastric volume over time were tested using linear mixed models, with time, treatment, and treatment*time interactions as fixed factors and baseline gastric volume as a covariate. Tukey HSD corrected post-hoc t-tests were used to compare individual time points.

Differences in plasma concentrations of FFA, glucose, insulin, serum NMR-based metabolites, and subjective ratings over time were tested by using linear mixed models, testing for main effects of time, treatment, and treatment*time interactions. Baseline values were added as covariate. False discovery Rate (FDR) correction was used for the NMR-based metabolites (Storey, 2002).

Normality of the data was confirmed with quantile-quantile (QQ) plots of the residuals. For insulin, FFA and prospective consumption a logarithmic transformation was applied to create a normal distribution. All statistical analyses were performed using the R statical software (version 4.0.2). The significance threshold was set at p = 0.05. Data are expressed as mean ± SE unless stated otherwise.

## 3 Results

### 3.1 In vitro digestion

To investigate emulsion stability of the EF and CF (**Table 1**) under gastric conditions, *in vitro* digestions were performed under both simulated infant and adult conditions (**Supplementary Figure 3**). Under both simulated infant and adult conditions, the EF displayed an earlier, i.e., at a higher pH, onset of layer formation compared to the CF. In addition, the formed lipid layer in the EF appeared to be more dense compared to the CF.

### 3.2 In vivo trial

#### 3.2.1 Gastric top layer formation

Onset time of the high-fat top layer, maximum top layer volume, and time to maximum top layer volume are described in **Table 2**. The onset of the top layer was 13.4 ± 5.1 min earlier for the EF compared to the CF (p = 0.017, 26.1 ± 4.4 min, and 39.5 ± 3.3 min for the EF and CF respectively) (**Supplementary Figure 4**). When baseline gastric volume was added as covariate the effect became more significant with a mean difference of 13.6 ± 3.6 min (p = 0.001). Time to maximum top layer volume was shorter and maximum top layer volume was lower for the EF (p = 0.022 and 0.033, respectively). Top layer AUC did not differ between treatments (p = 0.915).

**Table 2.** Differences in top layer formation between the two formulae (mean ± SE).

| Characteristic | Control | Experimental | Mean difference (95% CI) | P-value |
| --- | --- | --- | --- | --- |
| Onset time (min) | 39.5 ± 4.4 | 26.1 ± 3.3 | 13.4 (2.7 – 24.1) | 0.017 |
| Maximum top layer<br>volume (ml) | 203.4 ± 23.5 | 154.6 ± 10.9 | 49.0 (4.4 – 93.6) | 0.033 |
| Time to maximum top<br>layer volume (min) | 41.3 ± 4.7 | 27.4 ± 3.5 | 13.9 (2.3 – 25.7) | 0.022 |
| AUC (ml*min) | 5163 ± 642 | 5090 ± 444 | 72.7 (-1343 – 1488) | 0.915 |

Although the AUC was similar for both formulae, top layer volume over time showed a significant treatment by time interaction (p < 0.001) with lower volumes for the EF at all time points except t = 100.

The individual curves for top layer volume over time can be found in **supplementary figure 5**.

#### 3.2.2 Total gastric volume

Total gastric volume showed a linear decline over time for both formulae (**Figure 3**), with higher volumes for the EF (treatment effect, mean difference = 15.0 ml, p < 0.001). However, treatment by time interaction was not significant (p = 0.323). Total gastric volume AUC tended to be lower for the CF (p = 0.063).

**Figure 3.**
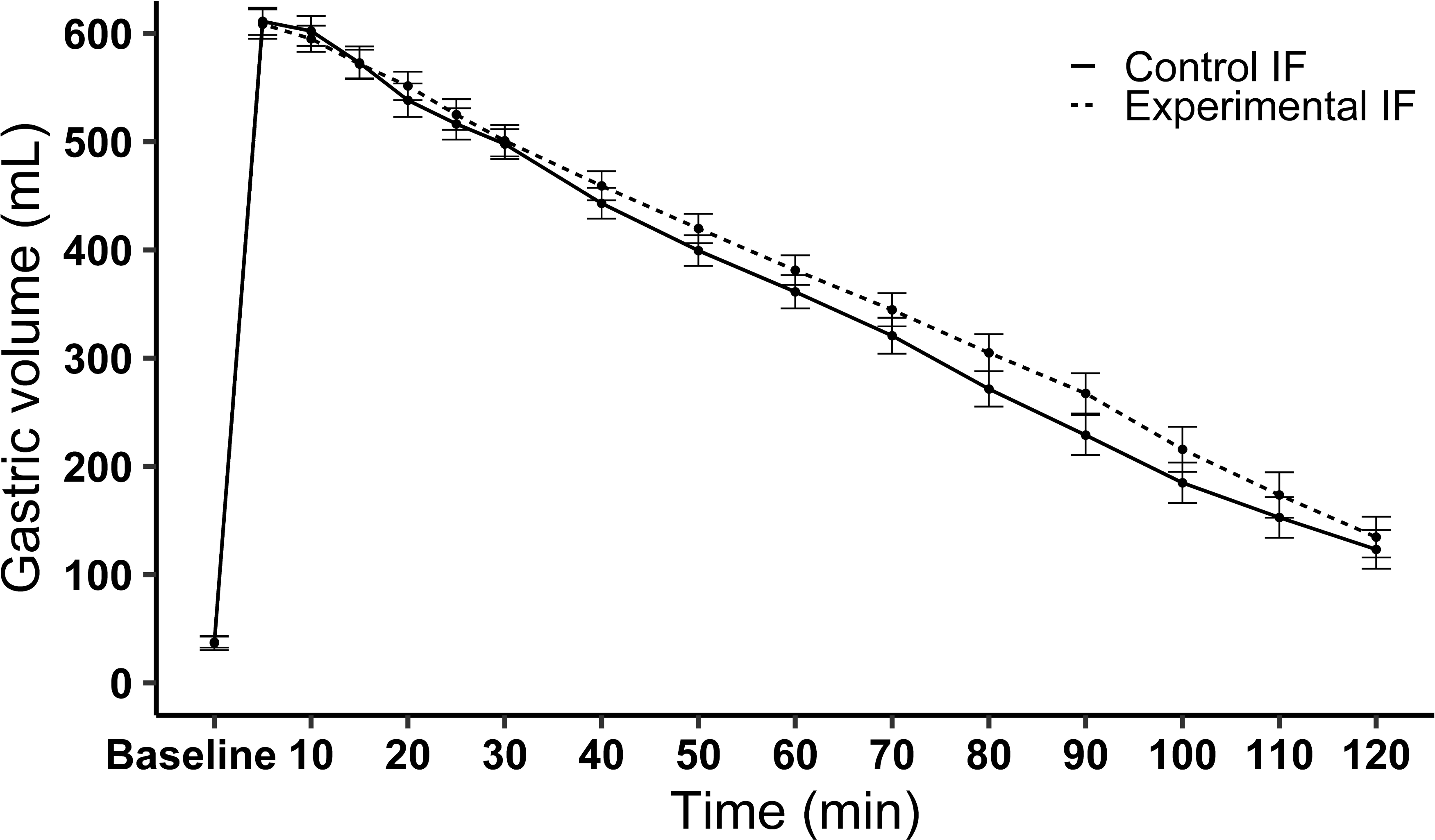
Mean ± SE total gastric volume over time of the two formulae.

#### 3.2.3 Correlations

Baseline gastric volume correlated with the top layer onset time for both the EF (r = - 0.82, p < 0.001) and the CF (r = −0.72, p < 0.001) (**Supplementary Figure 6**). Baseline gastric volume were also correlated with maximum top layer volume (r = 0.65 for EF, p < 0.001 and 0.46 for CF, p < 0.001) and time to maximum top layer (r = −0.80 for EF, p = 0.002 and r = −0.71 for CF, p = 0.046).

#### 3.2.4 Postprandial blood response

No treatment effects were found for postprandial plasma FFA and glucose (p = 0.763 and 0.325, respectively) (**Supplementary Figure 7)**. However, plasma insulin concentrations were lower for the EF (p = 0.040) (**Figure 4**). No treatment by time interaction was observed for these three parameters (p = 0.835, 0.881, and 0.547, respectively).

**Figure 4.**
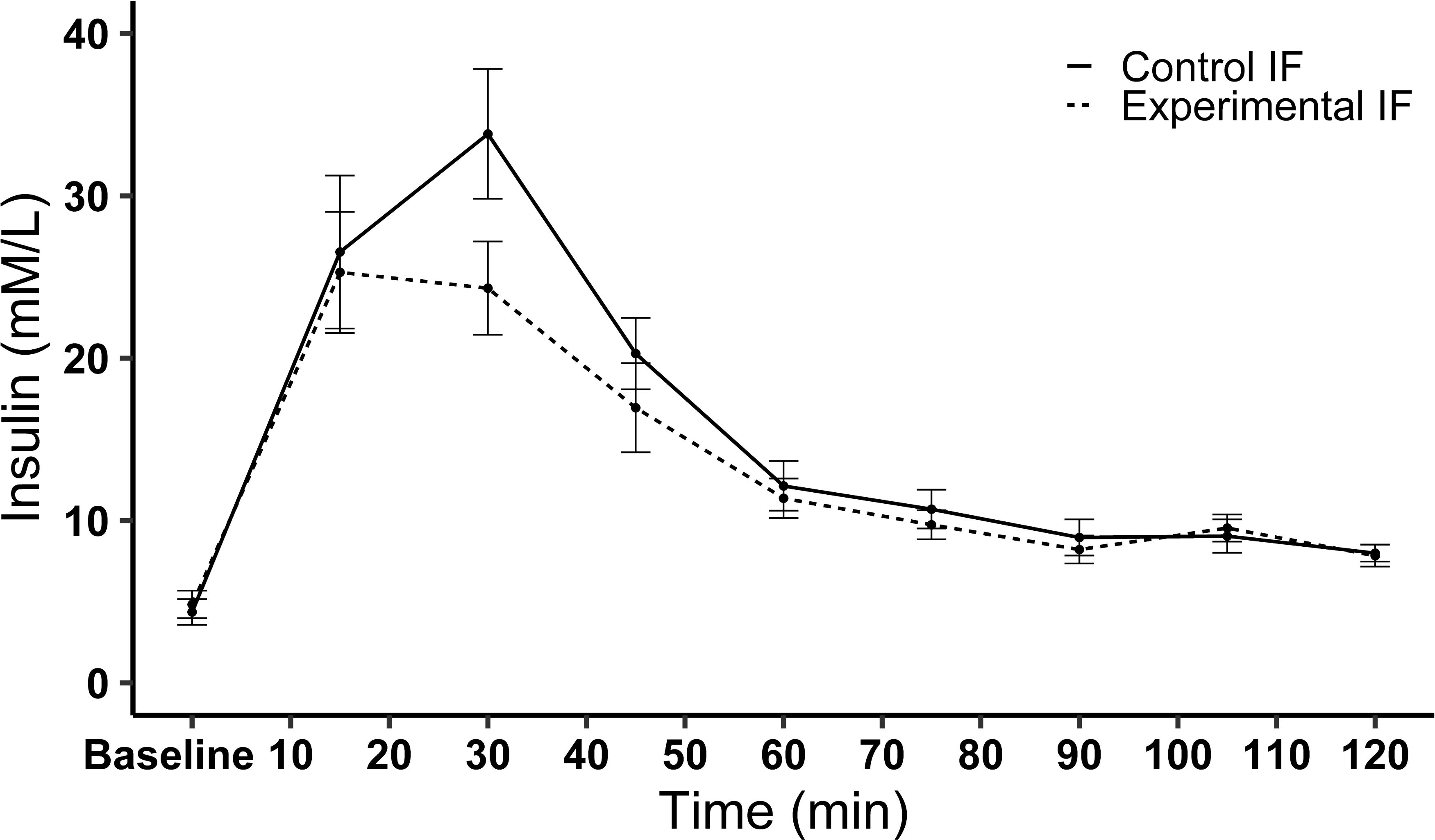
Mean ± SE plasma insulin concentrations over time after ingestion of the formulae.

Treatment effects were found for 63 of the NMR-based metabolites (**Supplementary Figure 8)**. Differences were mainly found in cholesterol- and phospholipid-related metabolites, which all had lower levels for the CF. None of the metabolites showed a significant treatment by time interaction.

#### 3.2.5 Appetite & nausea ratings

There were no significant treatment effects for hunger, fullness, prospective consumption, and thirst (p = 0.751, 0.340, 0.706, and 0.757 respectively). Desire to eat tended to be lower for the CF (p = 0.078). Nausea was overall rated lower for the EF (p = 0.023), although all scores remained around zero. None of the ratings showed a significant interaction between treatment and time (p = 0.820, 0.575, 0.257, 0.662, 0.718, and 0.890 respectively). The graphs are shown in **Supplementary Figure 9**.

## 4 Discussion

This study compared gastric top layer formation, gastric emptying, and post-prandial plasma metabolomics of an experimental minimally processed IF containing MFGM with that of a control IF. Onset of the high-fat gastric top layer was earlier for the EF and top layer volume was lower compared to the CF. For both formulae, total gastric volume emptied almost linear over time, with a higher volume for the EF. No treatment differences were found in post-prandial plasma concentrations of FFA, insulin, and glucose. NMR-based metabolomics mainly showed differences in postprandial serum phospholipid and cholesterol related metabolomics, with higher concentrations for the EF.

Gastric top layer volume over time was lower for the EF and the layer was formed ∼13 minutes sooner for the EF. This is in line with the observations in the *in vitro* digestion, which demonstrated layer formation at higher pH (i.e., in an earlier phase of gastric digestion) for the EF under both simulated infant and adult conditions. The lower gastric volumes for the EF are likely the result of the earlier top layer formation, resulting in a more even distribution over time. This earlier onset of emulsion destabilization is in line with our hypothesis and can most likely be explained by the mild processing. As a result of the lower heating compared to the CF, fewer whey proteins are denatured and complexed with caseins at the fat globule interface. This in turn causes the emulsion to be less stable under gastric conditions as caseins, resulting from pepsin-mediated hydrolysis, will coagulate quickly under gastric conditions (Anema et al., 2004; Fox et al., 2015; Guyomarc’h et al., 2009; Raikos, 2010). In the CF top layer formation occurred later, most likely as a result of a slower hydrolysis of the casein-whey complex that stabilizes the emulsion. A faster gastric lipid layer formation may potentially better mimic the phased-release of nutrients in the intestine overall as observed with breastfeeding where the gastrointestinal tract will initially be exposed to a relative low-fat fraction (foremilk) followed by exposure to a relatively high-fat fraction (hindmilk) (Daly et al., 1993; Forsum & Lonnerdal, 1979; Hytten, 1954; Kent et al., 2006; Saarela et al., 2005).

For both formulae, gastric emptying over time followed a linear pattern, with lower gastric volumes for the CF after 30 min. However, individual timepoints did not show a significant difference and the AUC did not significantly differ between treatments, indicating that the effect is small. Moreover, this small effect also did not result in significant differences for FFA and glucose. Therefore, it should be questioned whether this small treatment difference in gastric volume is clinically relevant.

Plasma insulin showed a higher initial postprandial peak for the CF. As total levels of carbohydrates were only slightly higher in the EF, these observations are likely explained by the insulinotropic effect of proteins (Rietman et al., 2014). Most likely, the slower destabilization of the emulsion for the CF led to a more protein-rich fraction initially reaching the intestine compared to the EF, thereby initially promoting insulin secretion more prominent. Surprisingly few data is available with respect to post-prandial insulin responses after consumption of human milk, likely resulting from ethical challenges of (serial) blood sampling in infants. Interestingly, an adult study comparing, amongst others, human milk with bovine milk, human milk displayed the lowest insulin response (Gunnerud et al., 2012).

As identified previously by Camps et al. (2021), baseline (fasted) gastric juice volume was correlated with top layer formation. Higher baseline gastric juice volume was strongly associated with an earlier onset of the top layer. This is likely explained by its low pH and the presence of pepsin. When there is more gastric juice, the gastric pH will initially be lower and more pepsin will be available, resulting in an overall quicker coalescence and creaming. Relative to Camps et al. (2021) (200 ml), a larger ingestion volume (600 ml) was chosen to minimize the influence of baseline gastric juice.

Nevertheless, a strong correlation between fasting gastric juice volume and subsequent top layer formation was also observed in the current study. This underscores the relevance of taking baseline gastric content volumes into consideration, also because these show large day-to-day and intra-individual variability (Grimm et al., 2018). Interestingly, one of the participants did not show any top layer formation for both formulae. Although this participant did not show abnormal baseline gastric juice volumes, it could have been that this participant had low pepsin activity. It is known that there are large interindividual variabilities in pepsin activity (Walther et al., 2019).

Post-prandial concentrations of phospholipid and cholesterol related metabolites were higher for the EF. Most likely, this can be explained by the addition of MFGM-enriched whey to this formula, which, as compared to normal whey, is enriched with phospholipids and cholesterol (Venkat et al., 2022). No treatment differences were found in post-prandial plasma FFA and glucose concentrations. This is likely due to the overall similar nutritional composition of the two formulae concerning fat and carbohydrate source. In addition, especially for the FFA, the measurement time was relatively short in the current study. Plasma FFA concentrations rise during fasting and drop as soon as food is ingested. This drop happens due to the meal-induced secretion of insulin, which suppresses intracellular lipase and thereby lowers the release of FFA into the circulation (Albrink & Neuwirth, 1960; Fielding, 2011; Lairon et al., 2007). This is in line with the plasma FFA concentration changes found in this study, which showed an initial decrease after ingestion of the IF. Therefore, it is often recommended to measure FFA concentrations for a longer period. For example, Lairon et al. (2007) recommend a period of 6-8 h.

One of the limitations of the study is the use of adults instead of infants because of ethical considerations associated with MRI. The gastrointestinal tract of infants is not yet completely developed and therefore differs from that of adults. Amongst others, the minimum gastric pH of infants is higher compared to that of adults (3 - 4 compared to 0.5 - 2.3) and less pepsin is secreted (Poquet & Wooster, 2016). Since the pH influences the activity of pepsin and gastric lipase, this impacts digestive processes in the stomach, among which the coalescence and creaming into a high-fat top layer. The higher pH in infant gastric conditions, would most likely result in a slower destabilization of the emulsion. However, as both formulae were measured in adults, it is expected that the differences between treatments will remain similar. Moreover, in the semi-dynamic *in vitro* digestion model the difference between both formulae were observed under both infant and adult digestion conditions. Combined, these results thus suggest that the differences in coalescence and creaming that were observed will also occur in infants.

MRI usually requires participants to be scanned in a supine position. Studies have shown that sitting in an upright position accelerates gastric emptying compared to a supine position (Jones et al., 2006; Spiegel et al., 2000). However, these effects are small and since participants were scanned in the same position for both treatments, we expect the differences between treatments to remain similar. Moreover, from the perspective of infant nutrition a supine position after feeding is realistic.

In conclusion, this study shows that an experimental minimally processed infant formula containing MFGM-enriched whey had an accelerated gastric creaming as compared to a control formula. No effects on overall nutrient absorption over time were found, except for the cholesterol- and phospholipid-related metabolites which can most likely be attributed to the presence of MFGM-enriched whey in the experimental formula. Although the overall physiological consequences remain to be identified, a faster high-fat gastric top layer formation of infant formulae may potentially better mimic the phased-release of nutrients in the intestine as observed with breastfeeding where the gastrointestinal tract will initially be exposed to a relative low-fat fraction (foremilk) followed by exposure to a relatively high-fat fraction (hindmilk).

## Data Availability

All data produced in the present study are available upon reasonable request to the authors

## Author contributions and acknowledgements

PS, RT and TL: conceptualization and methodology; JR: investigation; JR: data curation; JR: formal analysis; JR: Writing - Original Draft; RT, TL and PS: Writing - Review & Editing; JR: visualization; PS had primary responsibility for final content. All authors read and approved the final manuscript. This study was funded by FrieslandCampina and Reina Tjoelker and Tim Lambers are employed by FrieslandCampina. All other authors declare no conflicts of interest. We thank Romy de Haas, Natacha Benbernou, Caya Lindner and Joris de Jong for assisting with data collection collection and João Paulo for advice on statistical analysis. The use of the 3T MRI was made possible by WUR Shared Research Facilities.

## List of abbreviations

BM: Breast Milk
BMI: Body Mass Index
FDR: False Discovery Rate
FFA: Free Fatty Acids
IF: Infant Formula
MFGM: Milk Fat Globule Membrane
MRI: Magnetic Resonance Imaging
NMR: Nuclear Magnetic Resonance
SGF: Simulated Gastric Fluid

