## Supplementary materials for "Intragastric behavior of an experimental infant formula may better mimic intragastric behavior of human milk as compared to a control formula"

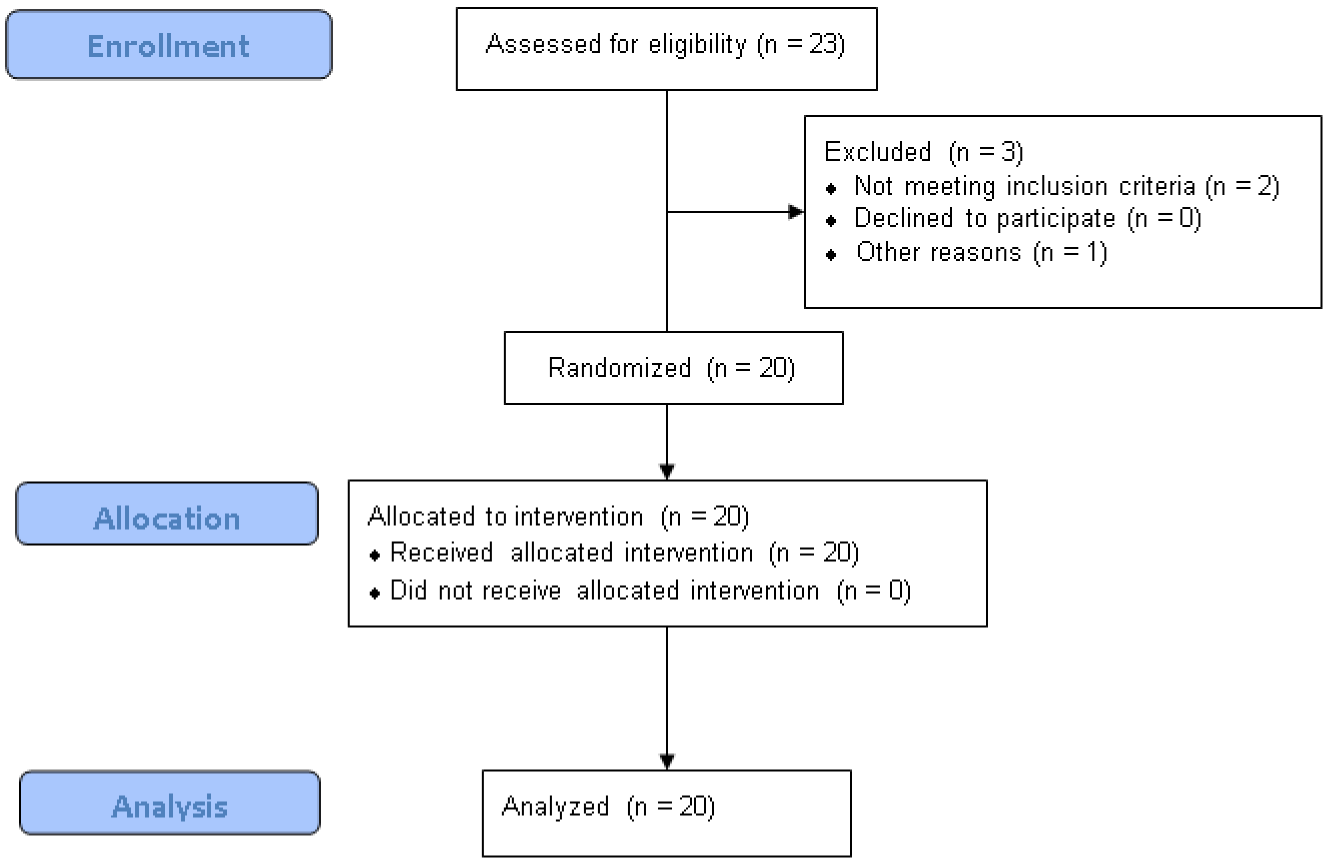


Supplementary Figure 1. Flow diagram for inclusion, treatment allocation and analysis.

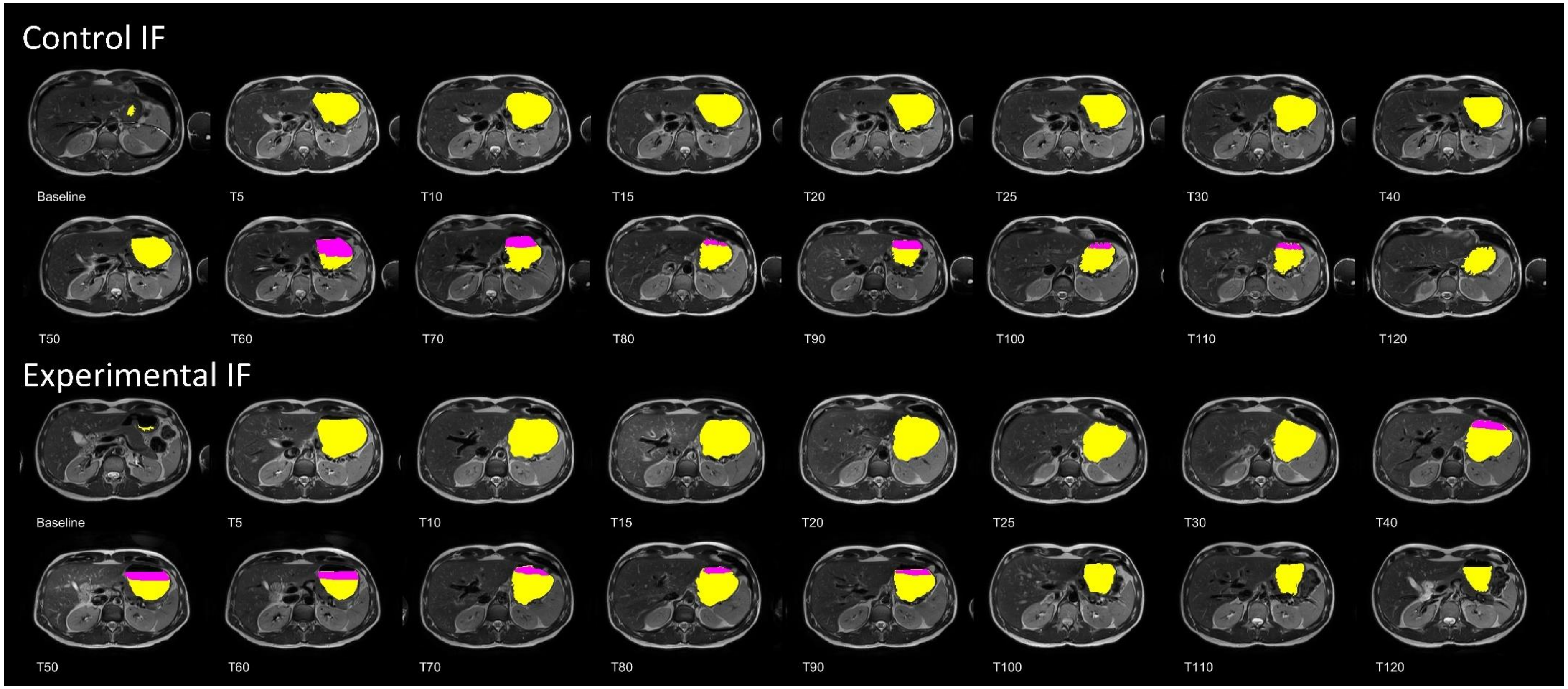


Supplementary Figure 2. Illustration of a stomach MRI time series from one participant with high-fat top layer volume (pink) and total gastric volume (yellow + pink) delineated for the CF and EF.

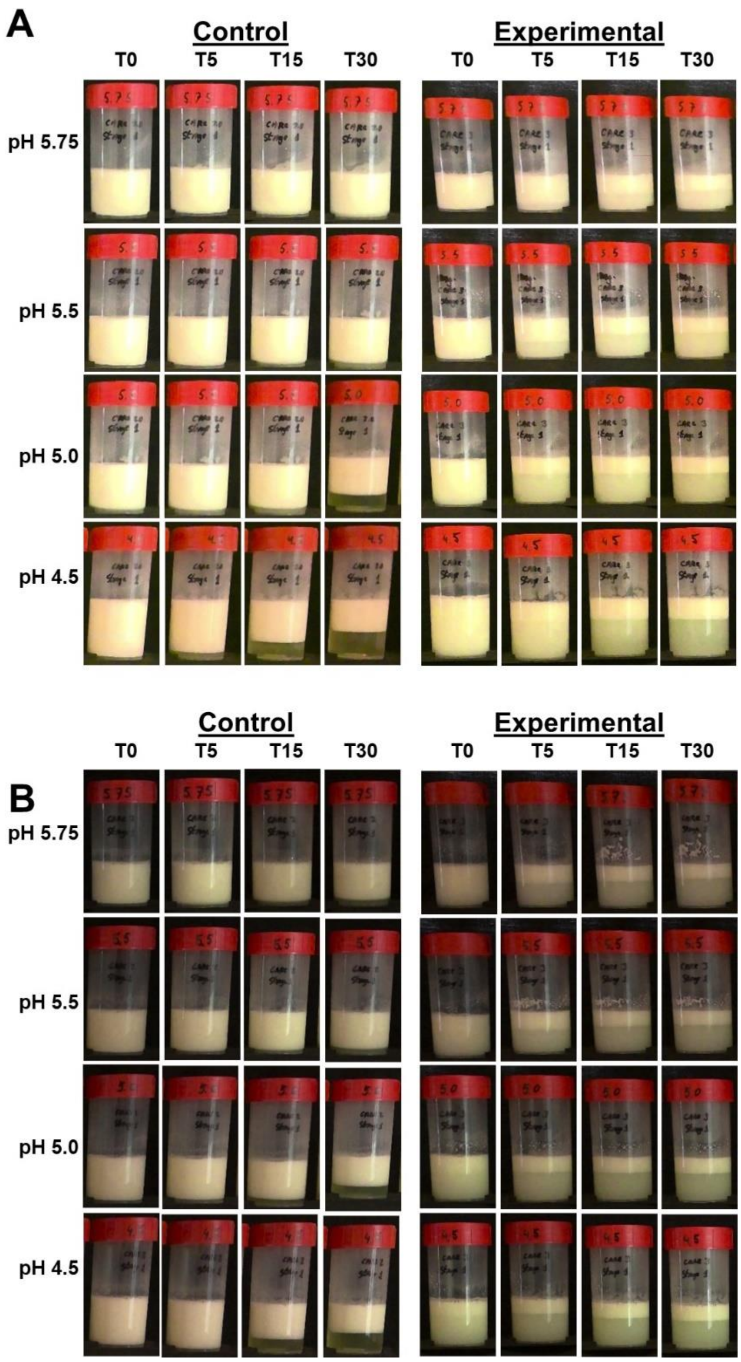


Supplementary Figure 3. Emulsion stability of EF and CF over time after *in vitro* digestion under simulated infant (A) and adult (B) conditions at different pH

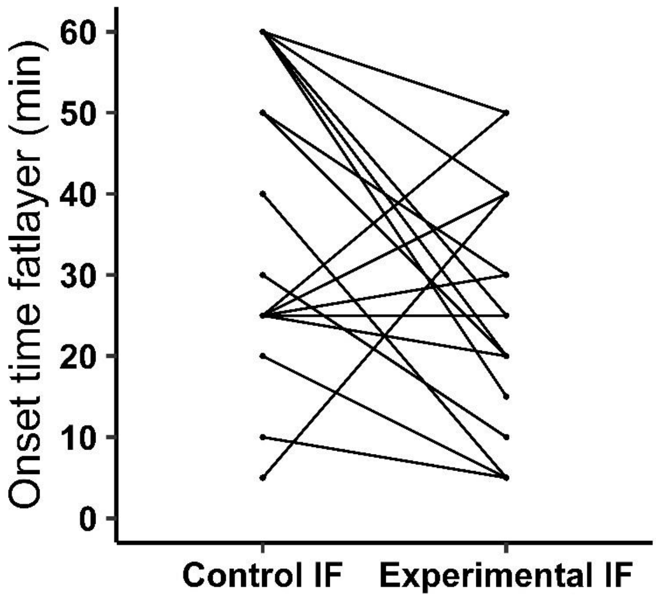


Supplementary Figure 4. Onset time of the gastric top layer (min) per participant for both formulae, CF and EF

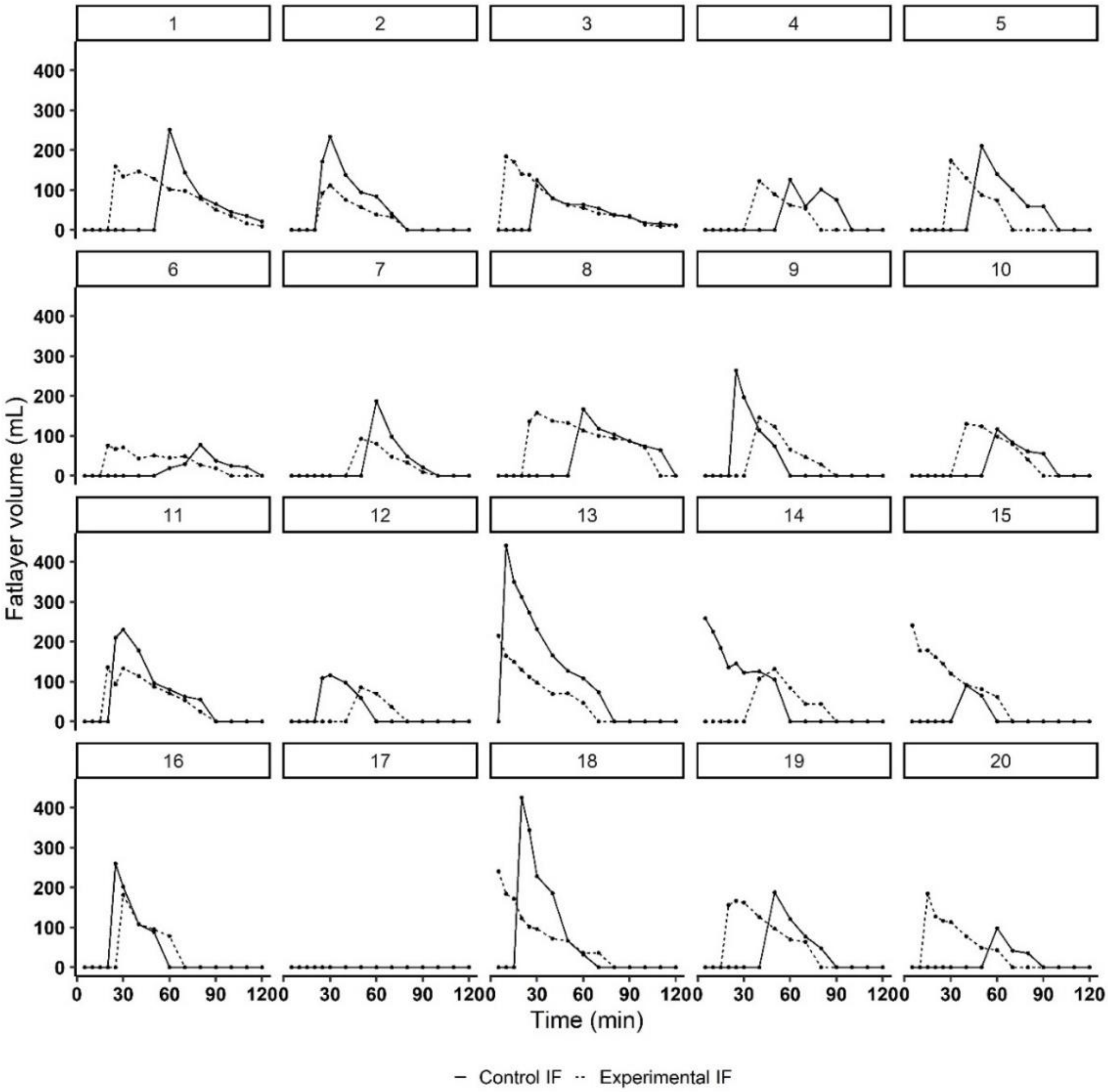


Supplementary Figure 5. Individual curves of the high-fat gastric top layer volume over time for CF and EF

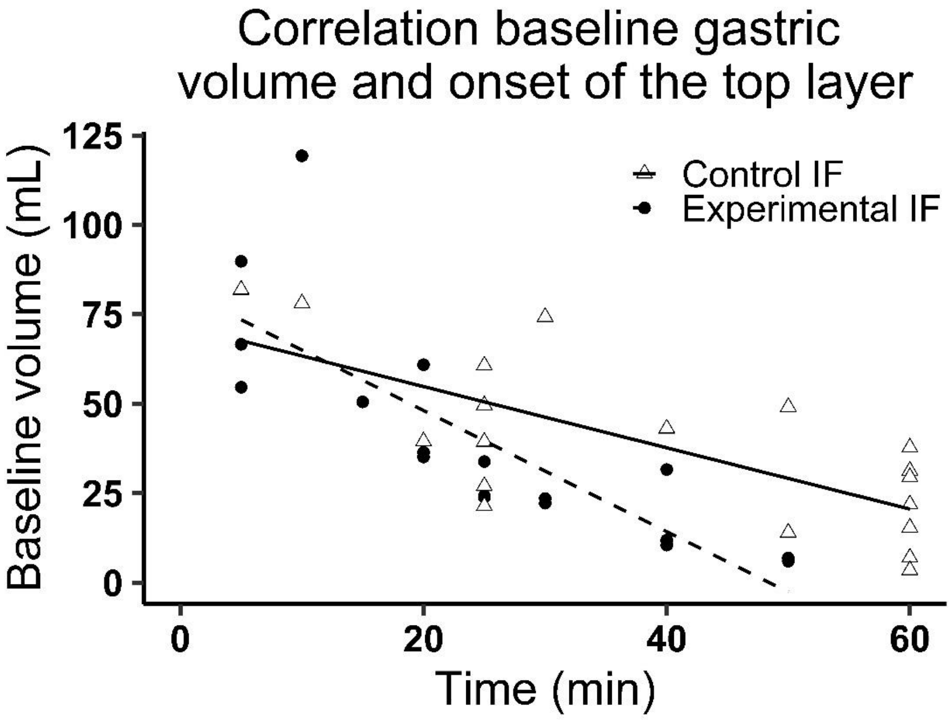


Supplementary Figure 6. Scatterplot of onset time of the gastric top layer (min) and baseline gastric volume of control IF (r = -0.72) and EF (r = -0.82).

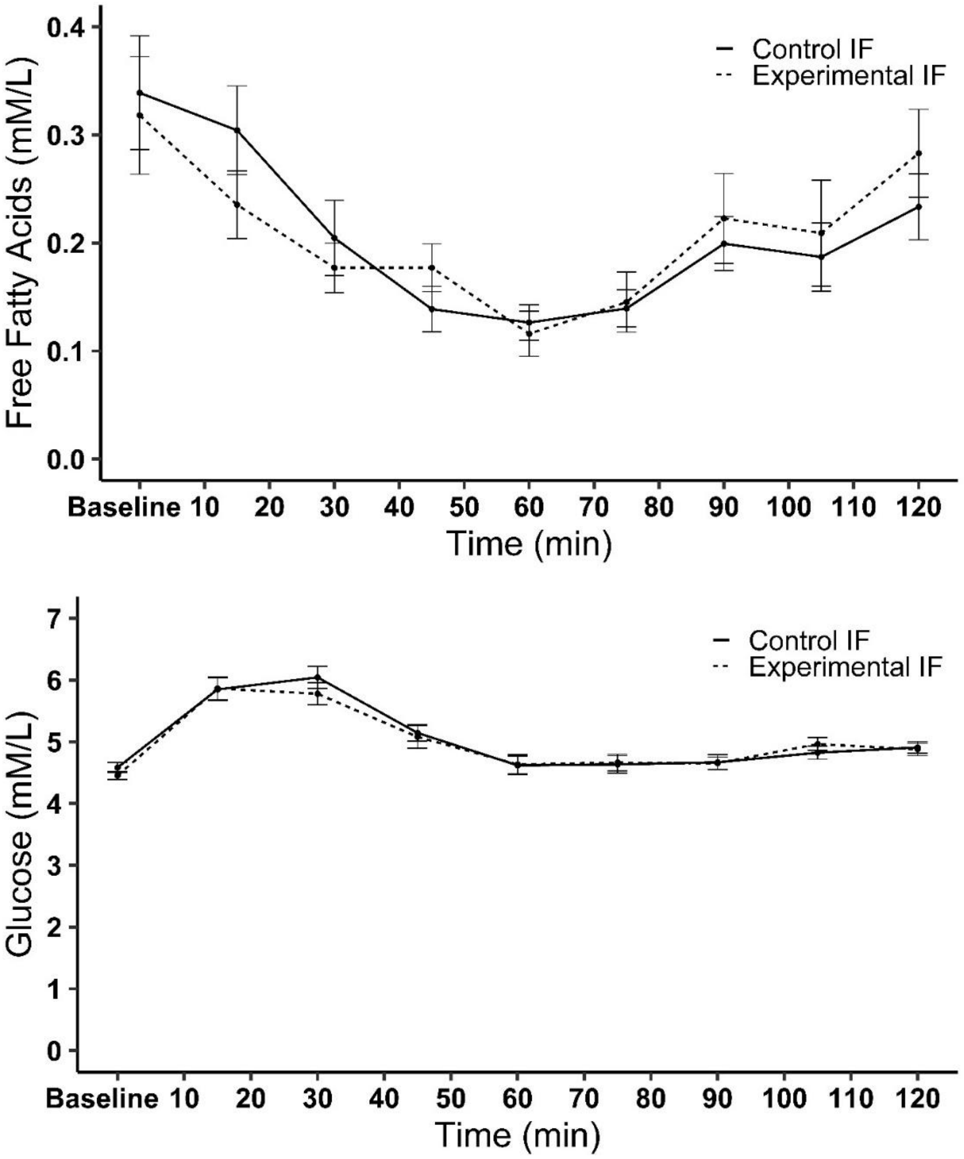


Supplementary Figure 7. Mean ± SE plasma FFA and glucose concentrations over time during the test session after ingestion of the formulae. No treatment effects were found for both postprandial FFA and glucose ($p = 0.763$ and 0.325, respectively). Moreover, there were no treatment by time interactions ($p = 0.835$ and 0.881, respectively).

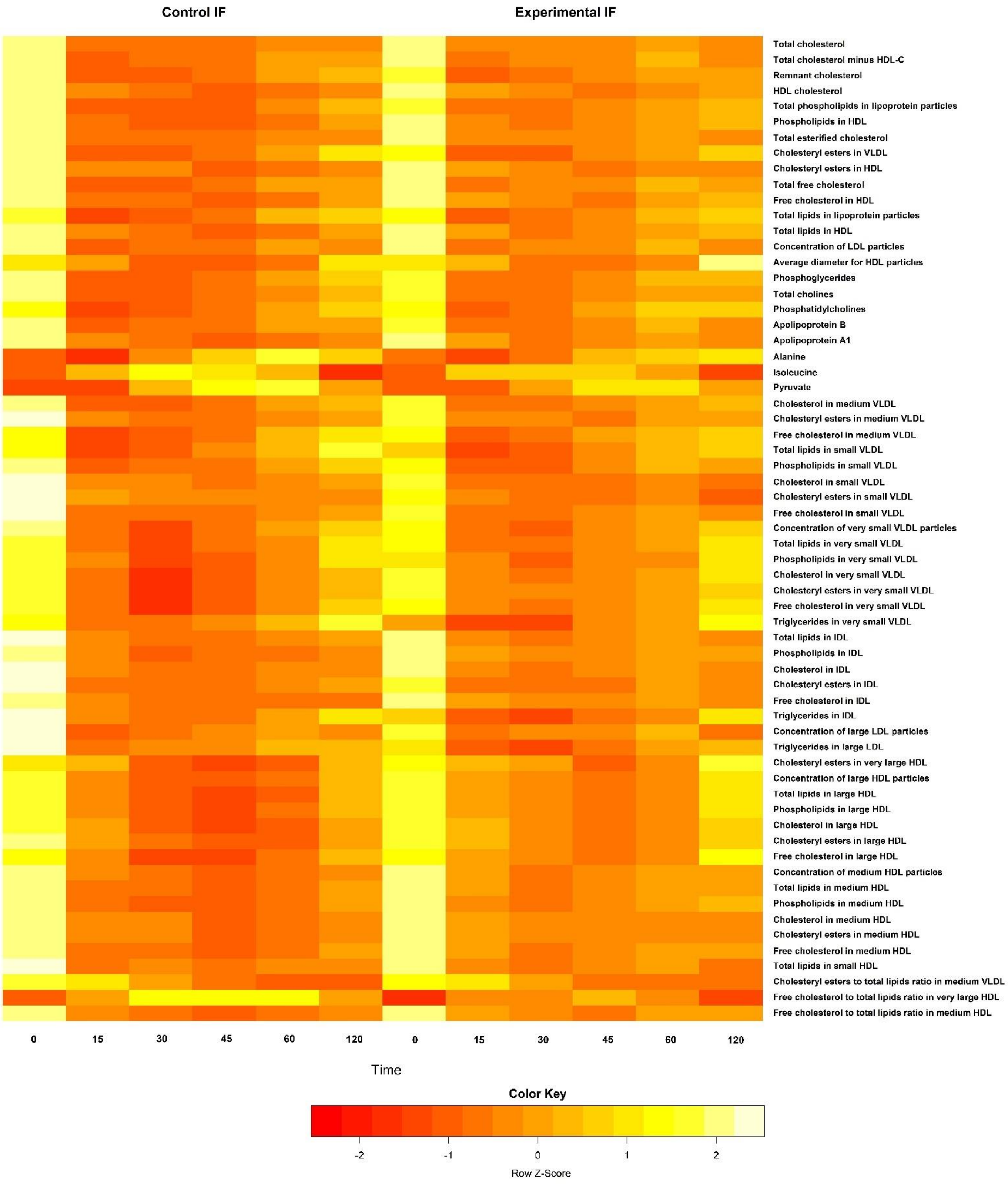


Supplementary Figure 8. NMR-based metabolomics over time that differ significantly ($p < 0.05$) between the control IF and EF. No significant effect of treatment by time was found.

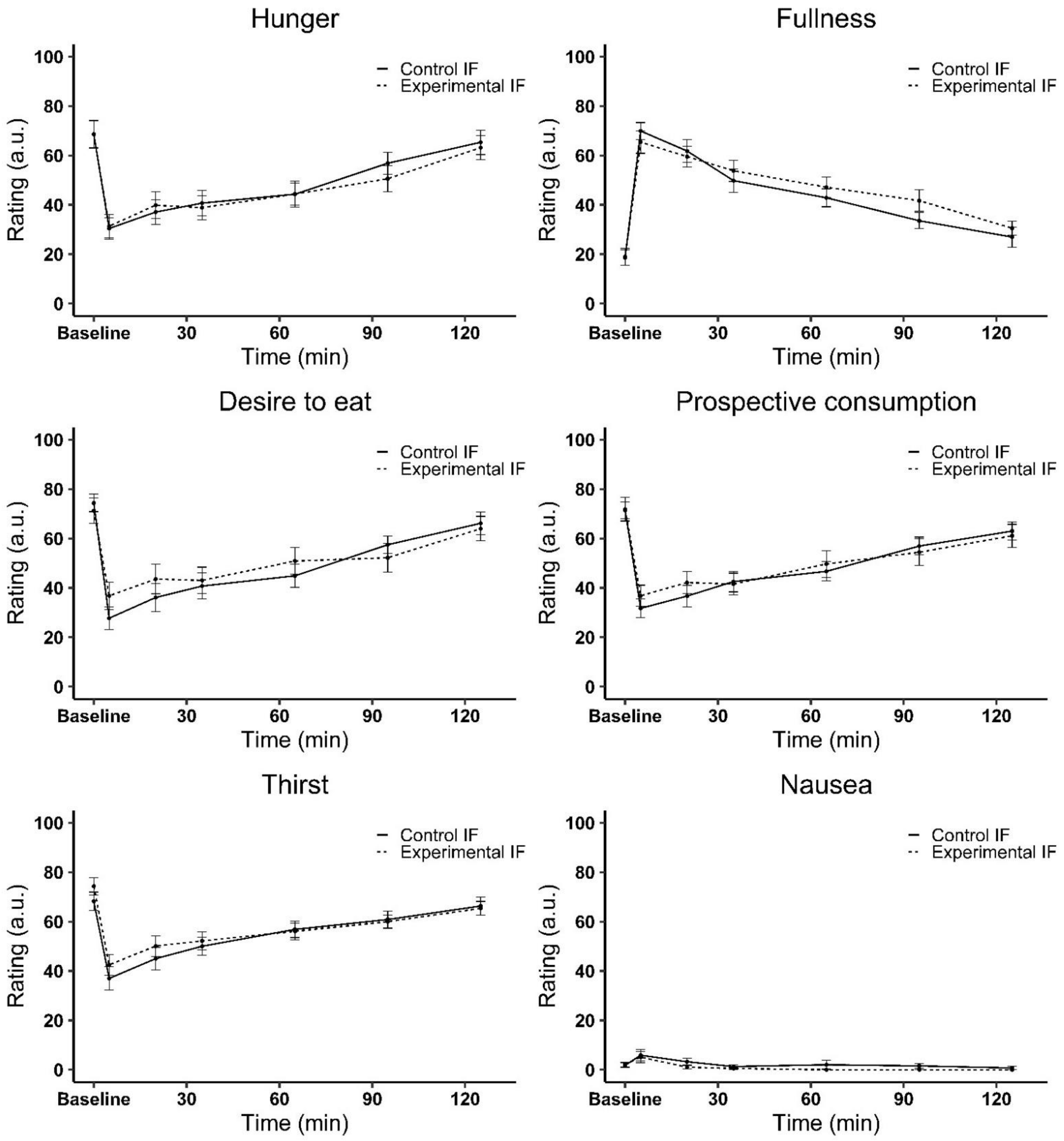


Supplementary Figure 9. Mean ± SE for appetite and nausea ratings over time per treatment. No significant differences were found for hunger, fullness, prospective consumption, and thirst ($p = 0.751$, 0.340, 0.706 and 0.757 respectively). Desire to eat tended to be lower for the control IF ($p = 0.078$). Nausea was overall rated lower for the EF ($p = 0.023$). None of the ratings showed a significant interaction between treatment and time ($p = 0.820$, 0.575, 0.257, 0.662, 0.718 and 0.890 respectively).
